# Mortality and outcomes after decompressive craniectomy in children with severe traumatic brain injury

**DOI:** 10.1101/2021.08.05.21261248

**Authors:** N. Bruns, O. Kamp, K. Lange, R. Lefering, U. Felderhoff-Müser, M. Dudda, C. Dohna-Schwake, TraumaRegister DGU

**Affiliations:** Department of Paediatrics I, Paediatric Intensive Care Medicine, University Hospital Essen, University of Duisburg-Essen, Essen, Germany; Department of Trauma, Hand and Reconstructive Surgery, University Hospital Essen, University of Duisburg-Essen, Essen, Germany; Faculty of Health, Institute for Research in Operative Medicine (IFOM), University of Witten/Herdecke, Cologne, Germany; Sektion Notfall-, Intensivmedizin und Schwerverletztenversorgung (Sektion NIS) of the German Trauma Society (Deutsche Gesellschaft für Unfallchirurgie)

**Author notes:** **Corresponding author:** Nora Bruns.

**Keywords:** Children, decompressive craniectomy, TraumaRegister DGU^®^, traumatic brain injury, outcomes

## Abstract

**Purpose:** The effect of decompressive craniectomy (DC) on mortality and outcomes in children with elevated intracranial pressure after severe head trauma is strongly debated and high-quality evidence is lacking. This study was conducted to determine whether DC in children with severe head trauma is associated with a decrease in mortality or poor outcomes at discharge from the intensive care unit.

**Methods:** Data on patients < 18 years of age treated in Germany, Austria, and Switzerland during a ten-year period were extracted from TraumaRegister DGU®, forming a retrospective multi-centre cohort study. Descriptive and multivariable analyses were performed to compare mortality and outcomes after decompressive craniectomy and medical management.

**Results:** 2507 patients were included, of which 402 underwent decompressive craniectomy. Mortality was 20.6 % in children undergoing DC compared to 13.7 % after medical management. Observed and predicted mortality after DC and medical management matched in all subgroups except in children between six and 17 years of age, where mortality after DC was lower than predicted. Poor outcome was observed in 27.6 % of DC patients vs. 16.1 % receiving medical management. Logistic regression revealed slightly negative effects of DC on mortality (odds ratio 1.20, not significant) and outcomes (odds ratio 1.56 (95% confidence interval 1.01-2.40).

**Conclusion:** DC did not decrease overall mortality or rates of poor outcome. However, children above six years of age may benefit from DC. High quality prospective studies are urgently needed.

## Introduction

Severe traumatic brain injury (TBI) is a relevant cause of morbidity and mortality in children around the globe [1]. Research interest in TBI has grown in recent years and progress has been made in gathering evidence on the best management of paediatric TBI [2]. The first tier of post-TBI treatment is designed to avoid exacerbation of cerebral injury by prevention or treatment of intracranial hypertension by optimising cerebral perfusion pressure and brain tissue oxygenation [3]. The second tier of therapy has a low evidence level and consists of rescue measures to control intracranial hypertension by application of hyperosmolar solutions, barbiturate infusion, hypothermia treatment, hyperventilation, and decompressive craniectomy [2, 3].

The potential of decompressive craniectomy (DC) to improve mortality and functional outcomes is highly debated. DC lowers elevated ICP in children [4-11], but one out of ten patients undergoing DC suffers from a complication requiring additional interventions [12]. The effect of DC on outcomes and mortality in children has not been studied in high quality randomised controlled trials (RCTs). In adults, two RCTs investigated the effect of DC on overall mortality and outcome [13, 14]. The studies differed in design, with the DECRA trial investigating the effect of DC on early and the RescueICP on late refractory ICP elevations. The findings supported DC for late rather than for early refractory ICP elevation with more survivors but more patients with low Glasgow Outcome Scores [13, 14].

In children, only one RCT including only 27 patients is available on DC [6], along with numerous small studies, case series, and one larger retrospective study including 150 children with DC [4-11, 15-19]. Studies on functional outcome reported better outcomes in children after DC compared to conservative treatment [4, 6, 20] or no difference [5]. Two studies found lower mortality in the DC groups compared to medical management [4, 20]. However, almost all reports are limited by small case numbers, retrospective and heterogenous study designs, and different outcome measures [3, 16]. Urgent questions concerning the timing and location of surgery, cut-off values for ICP, and the effect of DC on mortality and outcomes remain unanswered.

The aim of our study was to investigate the effect of DC on mortality and outcomes at discharge from the intensive care unit (ICU) in children with severe head trauma. To form a large international multi-centre cohort, we extracted data from a 10-year period from TraumaRegister DGU® (TR-DGU) from Germany, Austria, and Switzerland.

## Methods

### Data collection

The TraumaRegister DGU^®^ of the German Trauma Society (Deutsche Gesellschaft für Unfallchirurgie, DGU) was founded in 1993. The aim of this multi-centre database is a pseudonymised and standardised documentation of severely injured patients.

Data are collected prospectively in four consecutive time phases from the site of the accident until dis- charge from hospital: A) Pre-hospital phase, B) Emergency room and initial surgery, C) Intensive care unit and D) Discharge. The documentation includes detailed information on demographics, injury pattern, comorbidities, pre- and in-hospital management, course on intensive care unit, relevant laboratory findings including data on transfusion and outcome of each individual. The inclusion criterion is admission to hospital via emergency room with subsequent ICU/ICM care or reach the hospital with vital signs and die before admission to ICU.

The infrastructure for documentation, data management, and data analysis is provided by AUC - Academy for Trauma Surgery (AUC - Akademie der Unfallchirurgie GmbH), a company affiliated to the German Trauma Society. The scientific leadership is provided by the Committee on Emergency Medicine, Intensive Care and Trauma Management (Sektion NIS) of the German Trauma Society. The participating hospitals submit their data pseudonymised into a central database via a web-based application. Scientific data analysis is approved according to a peer review procedure laid down in the publication guideline of TraumaRegister DGU^®^. The participating hospitals are primarily located in Germany (90%), but a rising number of hospitals of other countries contribute data as well (at the moment from Austria, Belgium, Finland, Luxembourg, Slovenia, Switzerland, The Netherlands, and the United Arab Emirates). Currently, almost 30,000 cases from more than 650 hospitals are entered into the database per year.

Participation in TraumaRegister DGU^®^ is voluntary. For hospitals associated with TraumaNetzwerk DGU^®^, however, the entry of at least a basic data set is obligatory for reasons of quality assurance.

Patients aged < 18 years from Germany, Austria and Switzerland with Abbreviated Injury Scores (AIS) for the head ≥ 3 that were admitted between 2010 and 2019 (10 years) were included. Patients transferred to another hospital within the first 48 hours after admission were excluded due to missing data on final outcomes. Patients transferred in from another hospital were excluded as well. Only patient with complete standard documentation including surgical interventions were included.

### Measures of outcome and injury severity

Predicted mortality was calculated using the Revised Injury Severity Classification, version II (RISC II) score, which was developed and validated by the TR-DGU to predict survival of trauma patients based on 13 predictive factors [21]. Injury severity was measured using the Injury Severity Score (ISS) and Abbreviated Injury Score (AIS) for the head and body. Outcome was measured as good recovery, moderate disability, severe disability, vegetative state or death at ICU discharge.

We defined poor outcome as death or vegetative state. Good outcome was defined as severe disability, moderate disability, and good recovery.

### Guidelines and ethics approval

The present study is in line with the publication guidelines of the TR-DGU and is registered as project ID 2020-033. All methods concur with relevant guidelines and regulations and the study was approved by the ethics committee of the Medical Faculty of the University of Duisburg-Essen (21-10116-BO).

### Data analysis

Quantitative variables are presented as mean and standard deviation (SD) or median and interquartile range (IQR) in case of skewed data. For qualitative factors, absolute and relative frequencies are given. For mortality rates, 95 % confidence intervals (CI) were calculated for comparison of observed and predicted mortality by the RISC II score. Adjusted odds ratios with 95% CIs were calculated using a logistic regression model with hospital mortality as dependent variable. Covariates were selected based on previous knowledge on predictors of outcome in trauma patients. Cases with missing data were excluded from logistic regression.

Formal statistical testing was avoided due to the number of variables and large sample size. A statistical significance would have been found with a difference of more than ± 0.1 SD in case of a continuous measurement, and ± 3-5% in case of a categorical variable (depending on the prevalence).

Statistical analyses were performed with SPSS Version 26 (IBM Inc., Armonk, NY, USA) and SAS Enterprise Guide 7.1 (SAS Institute Inc., Cary, NC, USA)). Figures were produced with SAS Enterprise Guide 7.1.

## Results

### Description of the cohort

2507 patients met the inclusion criteria for the study. More than 90% of patients were treated in a Level 1 trauma centre and male sex prevailed (Table 1). DC was performed in 402 (16%) patients. Patients undergoing DC did not differ regarding age (Figure 1), but suffered more severe head injury, had higher ISS, lower initial Glasgow Coma Scale (GCS) values, and longer ICU and hospital stays compared to medical management (Table 1). In patients with AIS head 5, the findings on ISS and GCS were inverted with lower ISS and higher GCS values in patients receiving DC (Table 2).

**Table 1:**
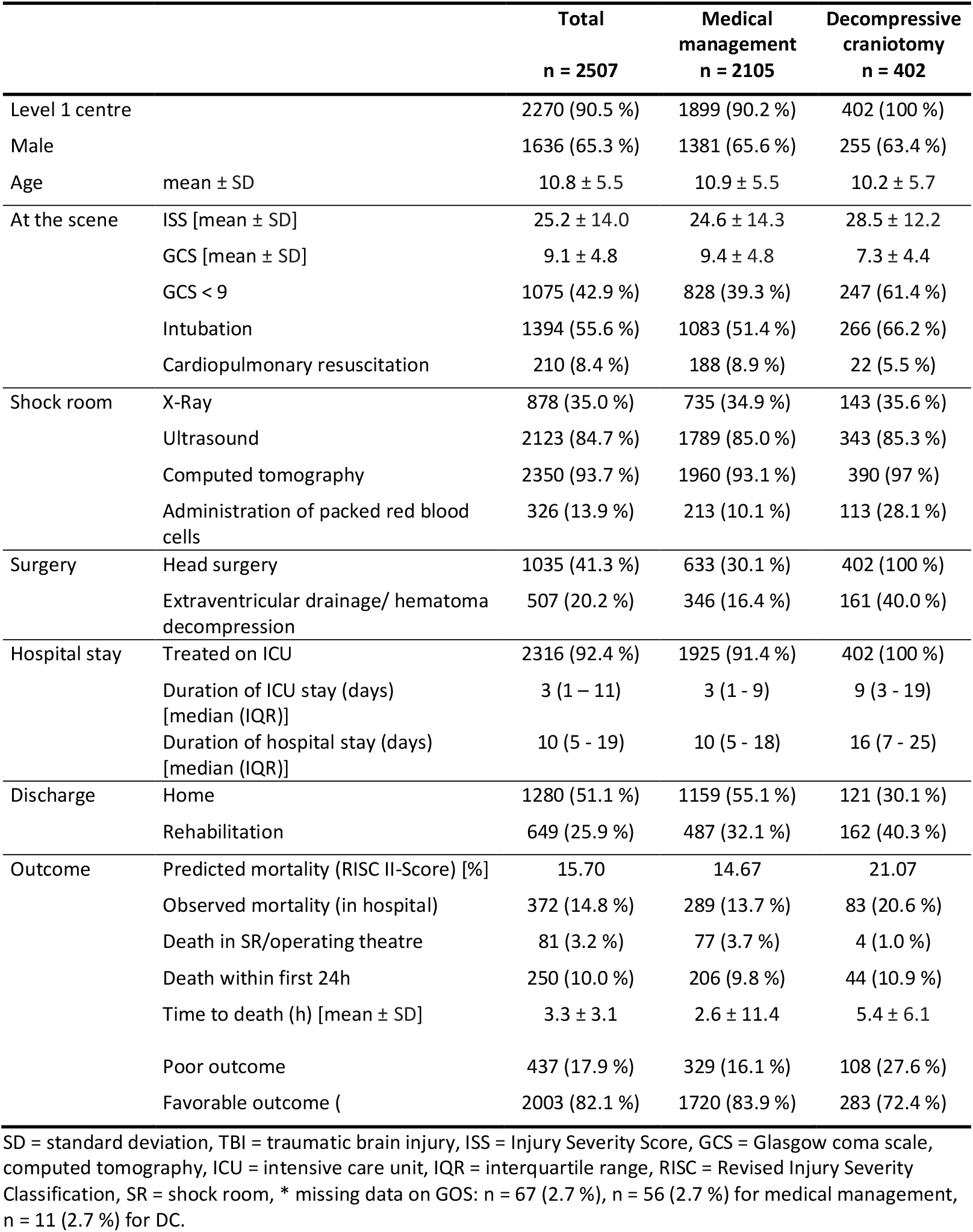
Patient characteristics

**Table 2:** Injury severity and combined outcomes by AIS head and treatment group

|  |  | AIS head 3 |  | AIS head 4 |  | AIS head 5 |  | AIS head 6 |  |
| --- | --- | --- | --- | --- | --- | --- | --- | --- | --- |
| N |  | 950 |  | 931 |  | 591 |  | 35 |  |
|  |  | MM | DC | MM | DC | MM | DC | MM | DC |
| N |  | 914 | 36 | 758 | 173 | 399 | 192 | 34 | 1 |
| ISS | mean ± SD | 17.2 ± 9.6 | 16.2 ± 6.9 | 24.3 ± 9.6 | 23.8 ± 9.6 | 38.0 ± 12.7 | 34.9 ± 11.0 | 75.0 ± 0.0 | 75.0 |
| GCS | mean ± SD | 11.2 ± 4.2 | 8.3 ± 4.9 | 9.6 ± 4.5 | 8.1 ± 4.5 | 5.7 ± 3.9 | 6.4 ± 4.1 | 3.3 ± 1.0 | 7.0 |
|  | median | 13 | 7 | 10 | 7 | 3 | 5 | 3 | 7 |
| Hospital stay<br>[days], all patients | median | 9 | 18 | 12 | 17 | 9 | 13 | 1 | 1 |
|  | IQR range | 5-16 | 10-25 | 7-19 | 10-25 | 2-21 | 4-26 | 1-2 | - |
| Hospital stay<br>[days], survivors | median | 9 | 20 | 12 | 19 | 20 | 20 | - | - |
|  | IQR range | 5-16 | 12-26 | 7-20 | 13-26 | 12-27 | 11-31 | - | - |
| Combined outcomes |  |  |  |  |  |  |  |  |  |
| Poor | N | 34 | 4 | 65 | 28 | 196 | 75 | 34 | 1 |
|  | % | 3.8 | 12.9 | 8.8 | 16.5 | 50.5 | 39.7 | 100.0 | 100.0 |
| Favorable | N | 852 | 27 | 676 | 142 | 192 | 114 | 0 | 0 |
|  | % | 96.2 | 87.1 | 91.2 | 83.5 | 49.5 | 60.3 | 0.0 | 0.0 |

AIS = abbreviated injury score, MM = medical management, DC = decompressive craniotomy, ISS = injury severity score, GCS = Glasgow coma scale, ICU = intensive care unit, SD = standard deviation. Note: Sums of absolute frequencies for combined outcomes can differ from the number *n* given in the headers due to missing data on GOS.

**Figure 1:**
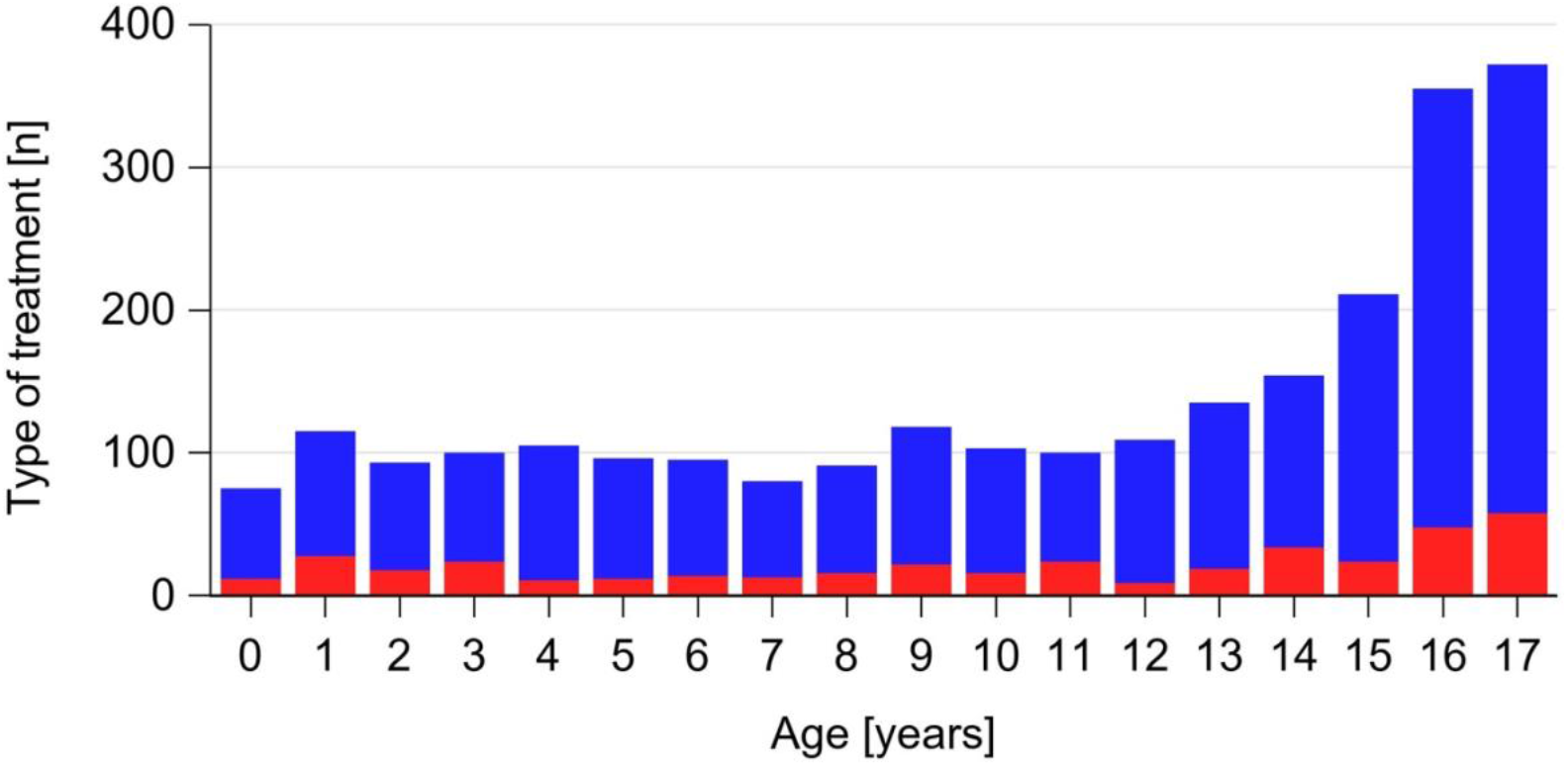
Frequencies of medical management (blue) and decompressive craniectomy (red) by age. The proportions of decompressive craniectomy versus medical management are constant, but absolute frequencies increase with age due to motor vehicle accidents.

Blunt trauma was the leading mechanism of injury and no differences were observed in the type of accident between groups (Table 2). Intracranial haemorrhage and brain edema were more prevalent in children undergoing decompressive craniectomy, while skull fractures and cerebral contusions occurring similarly often (Table 3).

**Table 3:** Mechanism and type of injury

|  |  | **Total**<br>**n = 2507** | **Medical**<br>**management**<br>**n = 2105** | **Decompressive**<br>**craniectomy**<br>**n = 402** |
| --- | --- | --- | --- | --- |
| Type of accident | Traffic | 1413 (56.3%) | 1183 (56.2%) | 230 (57.2%) |
|  | High fall | 408 (16.3%) | 347 (16.5%) | 61 (15.2%) |
|  | Low fall | 326 (13.0%) | 277 (13.2%) | 49 (12.2%) |
| Blunt trauma |  | 2362 (94.2%) | 1979 (94.0%) | 383 (95.2%) |
| TBI leading injury (AIS head > AIS rest) |  | 1977 (78.9%) | 1623 (77.1%) | 354 (88.1%) |
| Type of head injury | Contusion | 463 (18.4%) | 376 (17.9%) | 87 (21.6%) |
|  | Epidural haematoma | 345 (13.8%) | 221 (10.5%) | 114 (28.4%) |
|  | Subdural bleeding | 809 (32.3%) | 626 (29.7%) | 183 (45.5%) |
|  | Subarachnoidal bleeding | 214 (8.5%) | 154 (7.3%) | 60 (14.9%) |
|  | Intracranial bleeding | 285 (11.3%) | 213 (10.1%) | 72 (17.9%) |
|  | Brain edema | 424 (16.9%) | 298 (13.7%) | 126 (31.3%) |
|  | Brainstem injury | 103 (4.1%) | 93 (4.4%) | 10 (2.5%) |
|  | Skull fracture | 1237 (49.3%) | 1007 (47.8%) | 230 (57.2%) |

SD = standard deviation, TBI = traumatic brain injury, AIS = abbreviated injury score

### Mortality

Predicted and observed mortality was higher in children undergoing DC (Table 1) and with more severe head injury (Figure 2a + b). Mean time to death was 2.6 hours (± 3.1) after the accident for patients receiving medical management and 5.4 hours (± 11.4) for patients undergoing decompressive craniectomy. Mortality was higher in patients receiving medical management during the first 6 hours after the accident, but at 24 hours this finding was inverted (Figure 3).

**Figure 2:**
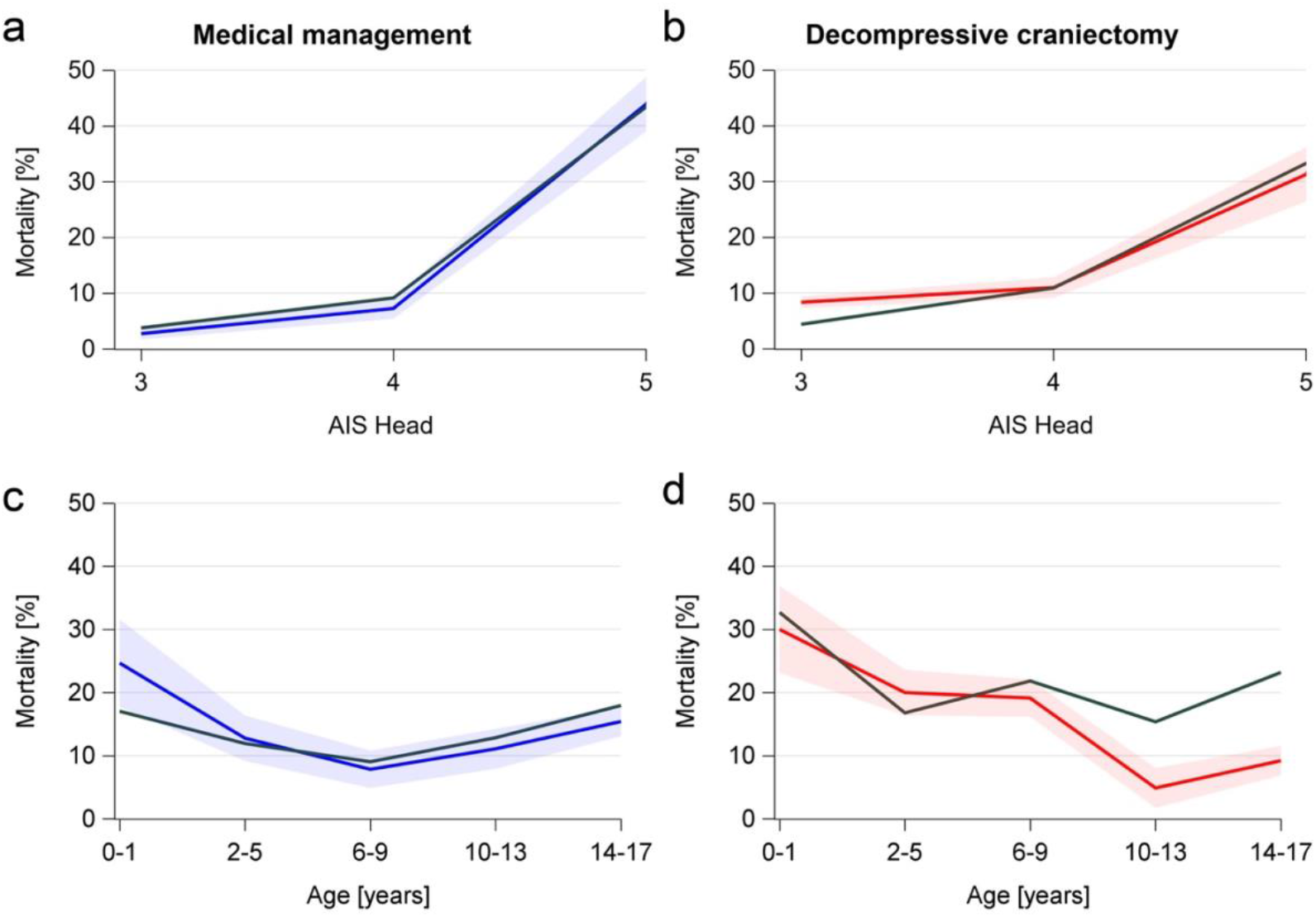
Observed mortality (grey line, based on RISC II score) and observed mortality (colored line) in subgroups of age (a+b) and severity of head injury (c+d). Colored bands: corresponding 95 % confidence intervals for the observed mortality. AIS = Abbreviated Injury Score.

**Figure 3:**
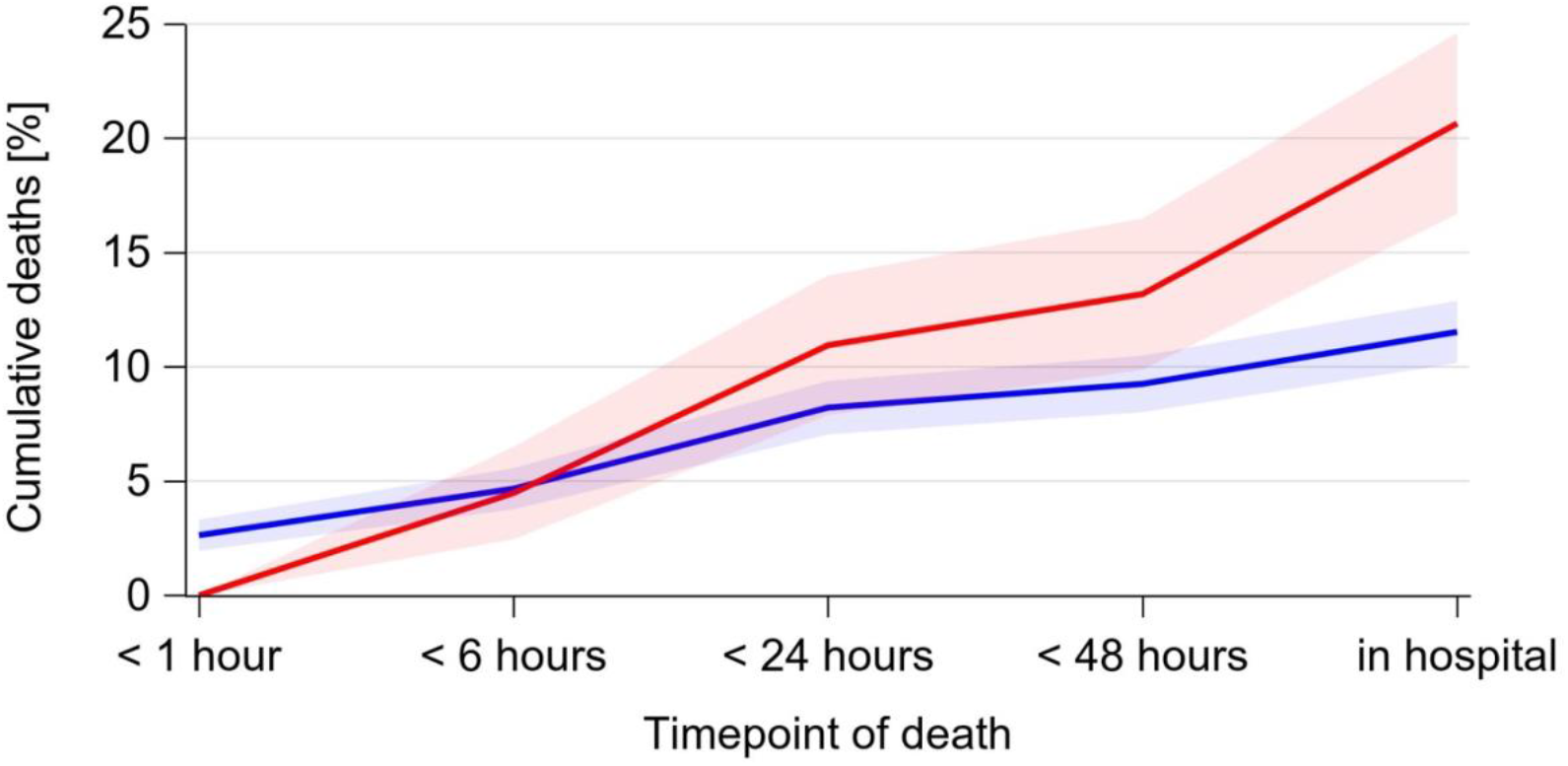
Cumulative deaths by time after the accident Blue line: medical management. Red line: decompressive craniectomy. Blue and red bands: corresponding 95 % confidence intervals.

Infants and adolescents had the highest predicted and observed mortality across groups and when receiving medical management (Figure 2c). In patients above 6 years of age undergoing decompressive craniectomy, the observed mortality was lower than predicted (Figure 2d).

### Combined outcomes

The percentage of favorable outcome (GOS 3-5) was higher in children with lower AIS head scores (Table 3, Figure 4). For AIS head 3 and 4, mortality and poor outcomes were more frequent in children undergoing DC compared to medical management, whereas in children with AIS head 5 lower mortality and better outcomes were observed after DC (Figures 2 + 4, Table 3).

**Figure 4:**
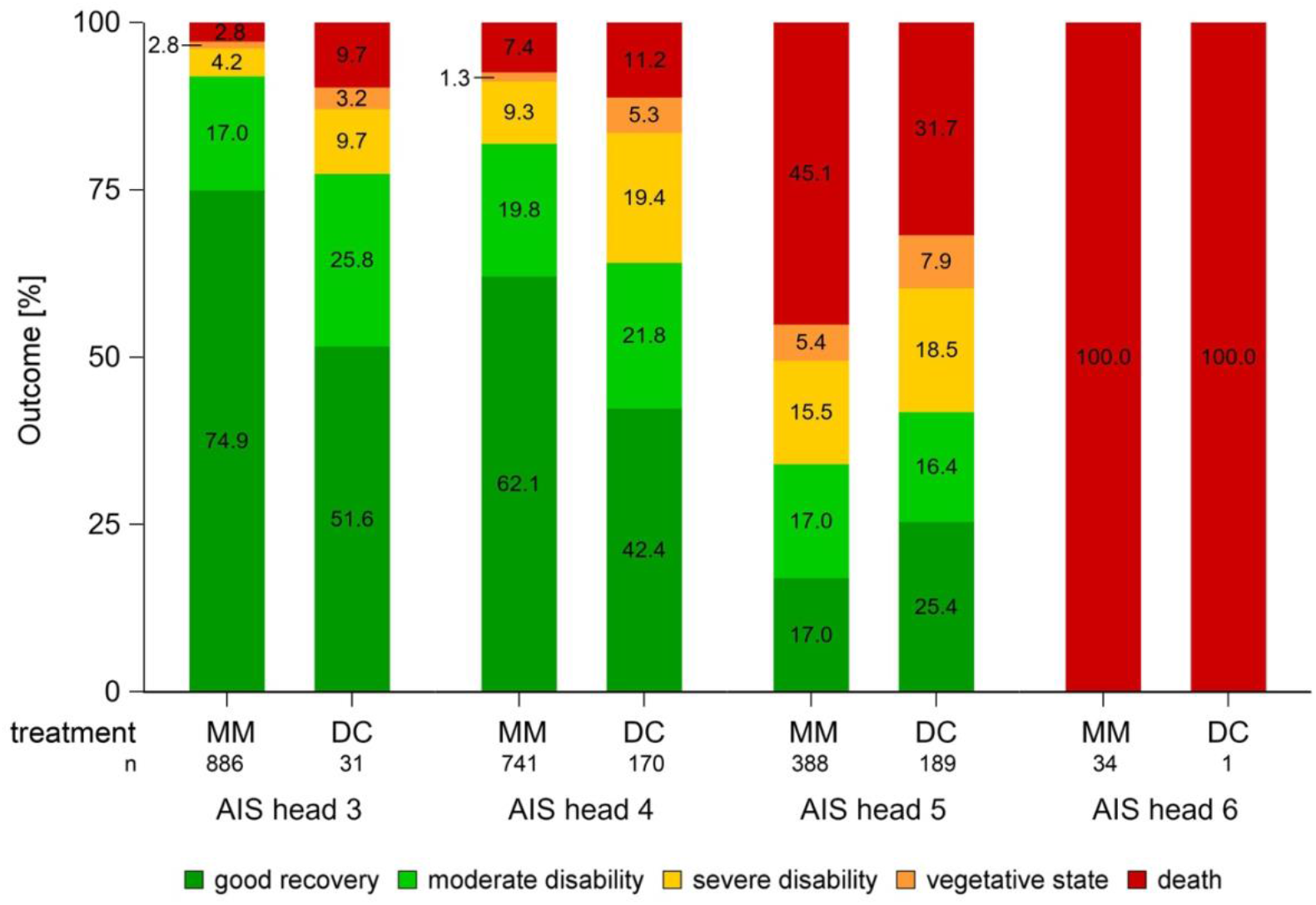
Mortality and outcomes by severity of head injury. MM = medical management, DC = decompressive craniectomy, AIS = Abbreviated Inury Score

### Multivariable analyses

Logistic regression was performed for patients with complete data sets (n = 2472, 98.6%). We found increased odds ratios for mortality (1.20, 95 % CI 0.74 – 1.95, not significant) and poor outcome (1.56, 95 % CI 1.01 – 2.40) in children undergoing DC (Figure 5). Children who received a cerebrospinal fluid (CSF) drain or evacuation of haemorrhage had lowered odds for mortality (OR 0.39, 95 % CI 0.25 – 0.63) and poor outcome (OR 0.57, 95 % CI 0.37 – 0.86). Brain edema and age below 6 years were independently associated with higher odds ratios for mortality and poor outcome. Intracerebral hemorrhage increased odds for poor outcomes. Epidural hematoma was independently associated with both lowered ORs for mortality and poor outcomes. Comprehensive results of the logistic regression are presented in Figure 5.

**Figure 5:**
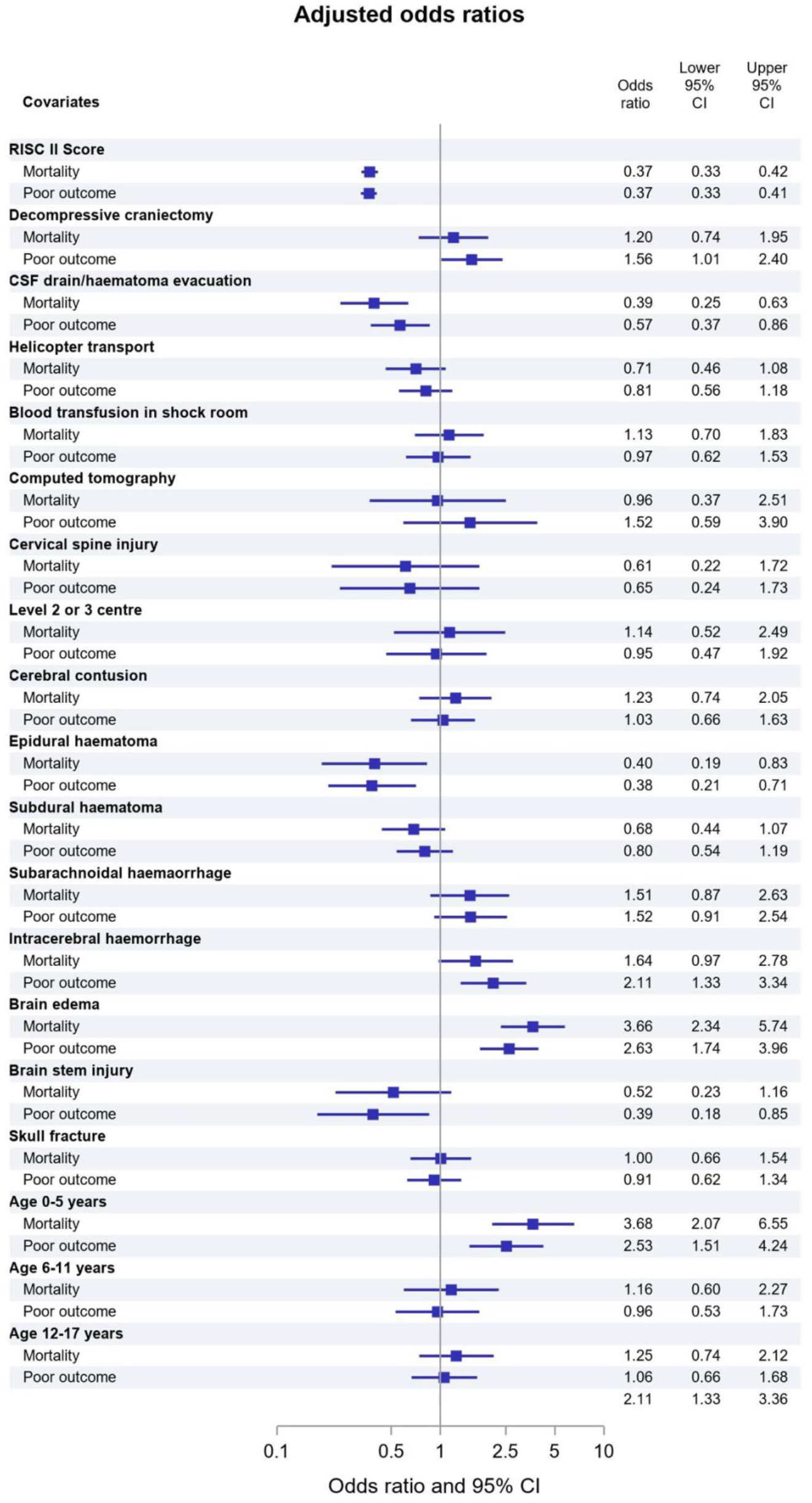
Adjusted odds ratios for mortality and poor outcomes. Squares: odds ratio, error bars: 95 % confidence intervals of the corresponding odds ratio. RISC = Revised Injury Severity Classification, CSF = cerebrospinal fluid drain. For optical convenience the scale of the x-axis is presented logarithmically.

## Discussion

This study on decompressive craniectomy in children with severe TBI is of high clinical relevance as the to date largest cohort of children with DC for TBI has been analysed. We found increased odds for death and poor outcome in children undergoing DC compared to medical management. Children receiving decompression by CSF drain or evacuation of hematoma had lower odds for death or poor outcome. Of note, in children above 6 years of age to 17 years mortality was lower when undergoing DC.

The age distribution in our study concurs with the typically described incidence peaks during infancy and adolescence for the occurrence of and mortality from TBI [1]: Additionally, the initial GCS values are in line with findings from other reports on children undergoing DC for severe TBI [22, 23]. Mortality was lower than predicted in children above six years of age undergoing decompressive craniectomy. Age below 6 years was an independent risk factor for death and poor outcome in the logistic regression. Manfiotto et al. reported that out of 150 children undergoing DC, those with poor outcome were significantly younger at the time of the accident [24]. While lower mortality after DC has been reported [4, 20], our study is the first to discriminate between age groups. According to Manfiotto’s and our study, the effect of DC on mortality may be less beneficial in infants and young children. Age-specific metabolic demands and susceptibility to tissue hypoxemia may contribute to the differences in outcomes.

Mortality and percentages of poor outcomes correlated with the severity of head trauma. In patients with AIS head 3 or 4 receiving DC, mortality and poor outcome were more frequent compared to patients with medical management. In patients with AIS head 5, this finding was inverted. However, observed mortality showed no deviation from predicted mortality in any of the AIS head subgroups. Several studies reported favorable outcomes several months or years after DC, with percentages varying between 50 and 100% [4, 7, 20, 23, 24]. In contrast, one study found that 38% of patients undergoing DC died and cognitive abnormalities were present in more than 50% of survivors [22]. The only paediatric RCT on this topic reported better outcomes after DC within 6 hours of ICP rise compared to medical management (54 vs. 14% good outcomes).

From all retrospective studies comparing DC and medical management groups, our study is the largest and the first to report possible negative effects of DC on mortality and outcomes. Two other studies point in a similar direction: Thomale et al. including 53 patients found no differences in functional outcomes between DC and medical management, and Kan et al. including 51 patients observed increased mortality when DC was performed raised intracranial pressure only [5, 9]. Further risk factors, such as generalised brain edema and the localisation of the lesion, likely play an important role in identifying patients who might benefit from DC. However, this has not yet been systematically investigated.

There are several limitations to our study. No data were available on the timing of DC, whether DC was performed as a primary or secondary intervention, and ICP values. This lack of information is a serious concern, because the RescueICP and DECRA studies suggest that timing of DC is important [13, 14]. As a consequence of the results from these two studies, the recommendations by the brain trauma foundation have been updated in favour of secondary, but not primary DC in adults [25]. From our study, no recommendations on the optimal timepoint of decompression nor on ICP values requiring DC can be given. A further limitation is the lack of information on long-term outcome. However, the strengths of our study are large data sets derived from an international multi-centre cohort that is continuously collected, updated, and subject to standardised documentation rules.

Although the overall level of evidence for practice recommendations remains low in children, DC is currently suggested to treat neurological deterioration, herniation or refractory intracranial hypertension (level III recommendation) [2, 3]. The aims are to control ICP, improve outcomes and reduce mortality [2, 3]. After adjustment for numerous clinical and predicting variables, results from this study do not support DC to reduce mortality and vegetative state, especially when compared to CSF drain or evacuation of hematoma. From our data it remains a matter of debate whether DC is either not the optimal treatment, possibly directly increases poor outcome or is simply a marker of clinical deterioration leading to such a rescue intervention. However, stratified analyses showed lower mortality than predicted in children above 6 years, indicating that DC might have positive effects in some paediatric patients.

Larger prospective trials are needed to determine the subgroups of paediatric patients who benefit from DC, tolerable ICP values, and the optimal timing and technique of surgery. DC needs to be compared not only to medical management, but also with surgical alternatives such as CSF drain or evacuation of hematoma. Unfortunately, sufficiently powered RCTs are difficult to conduct in this patient population. To our knowledge, the RANDECPED trial, designed to recruit 60 patients < 17 years and compare functional outcomes after 2 years between DC and medical management, is the only ongoing RCT on this topic. Despite the retrospective character of the current study, it has analysed by far the largest paediatric patient cohort. It adds relevant evidence in favour of decompression, but not clearly in favour of decompressive craniectomy. Our data call for careful and differentiated decision making when it comes to decompressive craniectomy in children.

## Conclusion

DC does not decrease mortality or rates of poor outcome in children with severe traumatic brain injury. Subgroups within the paediatric population may benefit from DC nonetheless, for example children above six years of age. Prospective studies including long-term neurological follow-up to identify subgroups and determine benefits are urgently needed to facilitate careful clinical decision making.

## Data Availability

The original data are the property of the AUC Akademie der Unfallchirurgie and were accessed via the TraumaRegister DGU®. Only aggregated data are available from the TraumaRegister DGU® for researchers who meet the criteria for access.

## Declarations

### Availability of data and material

The original data are the property of the AUC Akademie der Unfallchirurgie and were accessed via the TraumaRegister DGU^®^. Only aggregated data are available from the TraumaRegister DGU^®^ for researchers who meet the criteria for access.

### Code availability

Not applicable

### Authors’ contributions

Conceptualisation: NB, CDS, OK. Methodology: RL, OK, MD. Statistical analyses: RL. Interpretation of data: NB, CDS, RL, UFM. Writing – original draft preparation: NB. Writing – review and editing: CDS, RL, OK, UFM, MD.

### Ethics approval

The current study extracted data from the TraumaRegister DGU^®^ according to ethical guidelines. Ethics approval was obtained from the ethics committee of the Medical Faculty of the University of Duisburg-Essen (21-10116-BO).

### Consent to participate

Not applicable

### Consent for publication

All authors approved the final manuscript. The TraumaRegister DGU gave permission for publication.

### Take-home-message

Decompressive craniectomy after severe traumatic brain injury does not decrease mortality or poor outcome in children. Further studies are required to investigate whether subgroups might benefit from decompressive craniectomy, predominantly children older than six years.

